# Tocilizumab for patients with COVID-19 pneumonia. The TOCIVID-19 prospective phase 2 trial

**DOI:** 10.1101/2020.06.01.20119149

**Authors:** Francesco Perrone, Maria Carmela Piccirillo, Paolo Antonio Ascierto, Carlo Salvarani, Roberto Parrella, Anna Maria Marata, Patrizia Popoli, Laurenzia Ferraris, Massimiliano M. Marrocco-Trischitta, Diego Ripamonti, Francesca Binda, Paolo Bonfanti, Nicola Squillace, Francesco Castelli, Maria Lorenza Muiesan, Miriam Lichtner, Carlo Calzetti, Nicola Duccio Salerno, Luigi Atripaldi, Marco Cascella, Massimo Costantini, Giovanni Dolci, Nicola Cosimo Facciolongo, Fiorentino Fraganza, Marco Massari, Vincenzo Montesarchio, Cristina Mussini, Emanuele Alberto Negri, Gerardo Botti, Claudia Cardone, Piera Gargiulo, Adriano Gravina, Clorinda Schettino, Laura Arenare, Paolo Chiodini, Ciro Gallo, on behalf of the TOCIVID-19 investigators (see Appendix for a complete list), Italy

**Author notes:** contributed equally (Francesco Perrone, Maria Carmela Piccirillo). contributed equally (Paolo Chiodini, Ciro Gallo). **Corresponding author** Francesco Perrone, Clinical Trial Unit, Istituto Nazionale Tumori di Napoli.

## Abstract

**Background:** Tocilizumab blocks pro-inflammatory activity of interleukin-6 (IL-6), involved in pathogenesis of pneumonia the most frequent cause of death in COVID-19 patients.

**Methods:** A multicentre, single-arm, hypothesis-driven phase 2 trial was planned to study the effect of Tocilizumab on lethality rates at 14 and 30 days (co-primary endpoints). A cohort of patients consecutively enrolled after phase 2 was used as a validation dataset. A multivariable logistic regression was performed to generate hypotheses, while controlling for possible confounders.

**Results:** out of 301 patients in phase 2 intention-to-treat (ITT) analysis, 180 (59.8%) received tocilizumab. With 67 death events, lethality rates were 18.4% (97.5%CI: 13.6-24.0, P=0.52) and 22.4% (97.5%CI: 17.2-28.3, P<0.001) at 14 and 30 days. Lethality rates were lower in the validation dataset, including 920 patients. No signal of specific drug toxicity was reported. The multivariable logistic regression suggests tocilizumab might be more effective in patients not requiring mechanical respiratory support at baseline. Also, it supports a positive effect on lethality rate of the use of corticosteroids.

**Conclusions:** Tocilizumab reduced lethality rate at 30 days compared with null hypothesis, without significant toxicity. Such result support the use of tocilizumab while waiting for ongoing phase 3 trials.

**Registration:** EudraCT (2020-001110-38); clinicaltrials.gov (NCT04317092)

## Introduction

Pneumonia is the most frequent and serious complication of COVID-19, due to excessive host immune response causing an acute respiratory distress syndrome.[1-5]

Interleukin 6 (IL-6) is a pro-inflammatory cytokine implicated in several rheumatic diseases and in the so-called cytokine release syndrome (CRS). Tocilizumab is a recombinant humanized monoclonal antibody, directed against the IL-6 receptor. It is indicated for treating severe rheumatoid arthritis, systemic juvenile idiopathic polyarthritis and severe cytokine release syndrome (CRS) induced by chimeric antigen receptor T-cells (CAR-T).[6, 7]

Chinese researchers treated 21 patients with severe or critical COVID-19 pneumonia with tocilizumab 400 mg iv with efficacy in terms of reduction of oxygen requirement (15/20), resolution of radiologic lung lesions (19/21), normalization of lymphocyte count (10/19), and reduction of C-reactive protein levels (16/19).[8] These results prompted a randomised trial (tocilizumab vs control, ChiCTR2000029765).

On March 19^th^, 2020 during the ascending phase of the Italian breakout, we launched the TOCIVID-19 study, to describe efficacy of tocilizumab while controlling the highly increasing off-label use of the drug.

## Methods

TOCIVID-19, an academic multicentre clinical trial, was promoted by the National Cancer Institute of Naples and was approved for all Italian centres by the National Ethical Committee at the Lazzaro Spallanzani Institute on March 18^th^, 2020; two amendments followed on March 24^th^, 2020 and April 28^th^, 2020. TOCIVID-19 included a phase 2 study and a cohort study for patients not eligible for phase 2 or eligible but registered after the phase 2 sample size had been reached.[9] The study is coordinated through the web-based platform managed by the Clinical Trial Unit of the promoting centre.

### Selection of patients

Patients hospitalized due to clinical/instrumental signs of pneumonia, and with real-time PCR diagnosed SARS-CoV-2 infection, were eligible for the phase 2 study if they had oxygen saturation at rest in ambient air ≤93% or required oxygen support or mechanical ventilation either non-invasive or invasive (intubated less than 24 hours before registration). There was no limitation based on age and gender.

Patients were not eligible in case of known hypersensitivity to tocilizumab, known active infections or other clinical conditions that could not be treated or solved according to the judgment of the clinician and contraindicated tocilizumab, ALT/AST> 5 times the upper limit of the normality, neutrophils count <500/mmc, platelets <50.000/mmc, bowel diverticulitis or perforation.

Informed consent for participation in the study could be oral if a written consent was unfeasible. However, if patients lack capacity to consent due to disease severity, and an authorized representative was not immediately available, treatment could be administered by the treating physician on her/his own responsibility.

### Treatment

Tocilizumab was administered at the dose of 8 mg/kg up to a maximum of 800 mg per dose. Such dose is the same approved by FDA for the treatment of CRS following CAR-T therapy.[6] A second administration of tocilizumab (same dose) was allowed 12 hours after the first one if respiratory function had not recovered, at discretion of the Investigator.

Tocilizumab was supplied at no cost by Roche Italy. Due to the rapidly increasing request, a variable delay between the date of patient registration and drug availability at the clinical centres occurred. There was no contraindication for concomitant treatment of respiratory impairment; also, concomitant experimental antiviral treatment was allowed.

### Phase 2 study design and analysis

Sample size for the phase 2 study was initially calculated using 1-month lethality rate as the primary endpoint; based on March 10^th^ daily report on Italian breakout, 1-month mortality for the eligible population was estimated around 15%; 330 patients were planned to test the alternative hypothesis that tocilizumab may halve lethality rate (from 15% to 7.5%), with 99% power and 5% bilateral alpha error. The enrolment of 400 patients was planned to contrast possible ineligibility after registration. To increase the chance of producing useful data on time, the April 24^th^ amendment introduced 14-day lethality rate as co-primary endpoint. In addition, data accumulating between March 10^th^ and April 15^th^ clearly demonstrated a striking under-estimation of the 1-month lethality rate in the initial protocol. Therefore, expected lethality rates (null-hypotheses) at 14 and 30 days were redefined at 20% and 35%, respectively, based on data received from the Italian National Institute of Health.[10] The April 24^th^ amendment was proposed before extracting mortality data from the database, not being aware of the number and timing of recorded deaths. The planned sample size remained unchanged since it still allowed 99% and 95% power to recognize 10% absolute reduction at 14 and 30 days, respectively, with a significance level of 2.5% for each co-primary endpoint.

Primary analysis was performed in the intention to treat population (ITT), defined as all patients enrolled; a secondary analysis was done in the modified ITT (mITT) population with patients who had received at least one dose of the study drug.

Statistical analysis is detailed elsewhere.[10] Briefly, differences between groups of baseline characteristics, collected at the time of registration, are assessed for categorical variables using χ² test and for continuous variables using Wilcoxon rank-sum test. Patients discharged to home or low-intensity care setting are considered alive at the end-date of the follow-up period of 30 days. Exact 97.5% Clopper-Pearson confidence intervals (CI) are calculated for the proportions of death at 14 and 30 days. Pre-specified null hypotheses at days 14 and 30 are tested by a two-sided binomial test with alpha level equal to 0.025. Efficacy outcomes (with exact 95% CI) are described in baseline subgroups defined by demographics and clinical variables and compared with exact χ² test. Analyses were carried out using Stata version 14.0 (Stata Corp. College Station, TX, USA) and R version 3.6.1 (R Foundation for Statistical Computing, Vienna, Austria).

### Validation cohort

Patients eligible for phase 2 but exceeding the planned sample size, prospectively registered soon after the end of phase 2 enrolment, were considered as a validation cohort to possibly corroborate phase 2 findings. The same analyses performed in phase 2 were also done in the validation cohort. For the sake of efficiency, the results of the validation cohort are reported side by side those of phase 2.

### Joint cohort for safety analysis

Analysis of safety was performed joining phase 2 and validation cohorts and was limited to patients who received at least one dose of the study drug. Adverse events recorded from registration up to 30 days were graded according to CTCAE term (Version 5.0) and reported for each category and term as the worst grade suffered by patients through the whole period of observation after treatment administration.

### Hypothesis-generating multivariable analysis

Following the analysis of phase 2 and validation cohorts, multivariable logistic analysis, for each co-primary endpoint, was done to minimize the effect of two emergent biases: immortal time and indication bias. To reduce immortal time bias, patients who received tocilizumab four or more days after registration were excluded from analysis. To adjust for indication bias, that concerned the selection of patients to treat in the light of the delayed availability of drug, age (≤60, 61-70, >70), gender, type of respiratory support (oxygen, non-invasive mechanical ventilation [NIMV], invasive mechanical ventilation [IMV]), PaO2/FiO2 ratio (≤100, 101-200, >200, missing/not evaluated), population (phase 2 or validation) and geographical area (Lombardia, Veneto, Emilia-Romagna, other Northern regions, Centre, South and Islands) were entered as covariates in the multivariable logistic regression model. In addition, following the communication of positive results with dexametazone, treatment with corticosteroids (yes vs no) was added to the model. [11] Multivariable analysis was performed in the joint cohort because the number of events in the phase 2 population did not allow the adjustment for all the covariates. The interaction effects between treatment and the other covariates were tested in turn one at a time by Wald test and retained in the final model only if significant. Difference in the lethality rate between treated and untreated patients was calculated within specific subgroups and 95% CI was calculated by means of Agresti and Caffo method.[12] Description of such differences has to be considered as hypothesis generating only.

## Results

### Phase 2

From March 19^th^ (at 14:00) to March 20^th^ (at 12:45), 2020, 51 centres prospectively registered 402 patients for the phase 2 study (**Figure 1**, left side), of which 2 cases were duplicated and one case withdrew consent. In agreement with IDMC, 12 centres providing information on baseline characteristics and treatment for less than 25% of the enrolled patients were defined as un-cooperative and all the patients they had enrolled (either with or without missing data) were removed from the analysis. Therefore, the phase 2 ITT population include 301 patients. Out of these, 21 were found ineligible a posteriori (12 intubated more than 24 hours before registration, 7 registered after being already treated, 2 with both violations) but remained in the analysis. Geographical distribution and baseline characteristics of patients are summarized in **eFigure 1** (top graphs), **Table 1** (left side) and **eTables 1** to **3** (left side).

**Table 1.**
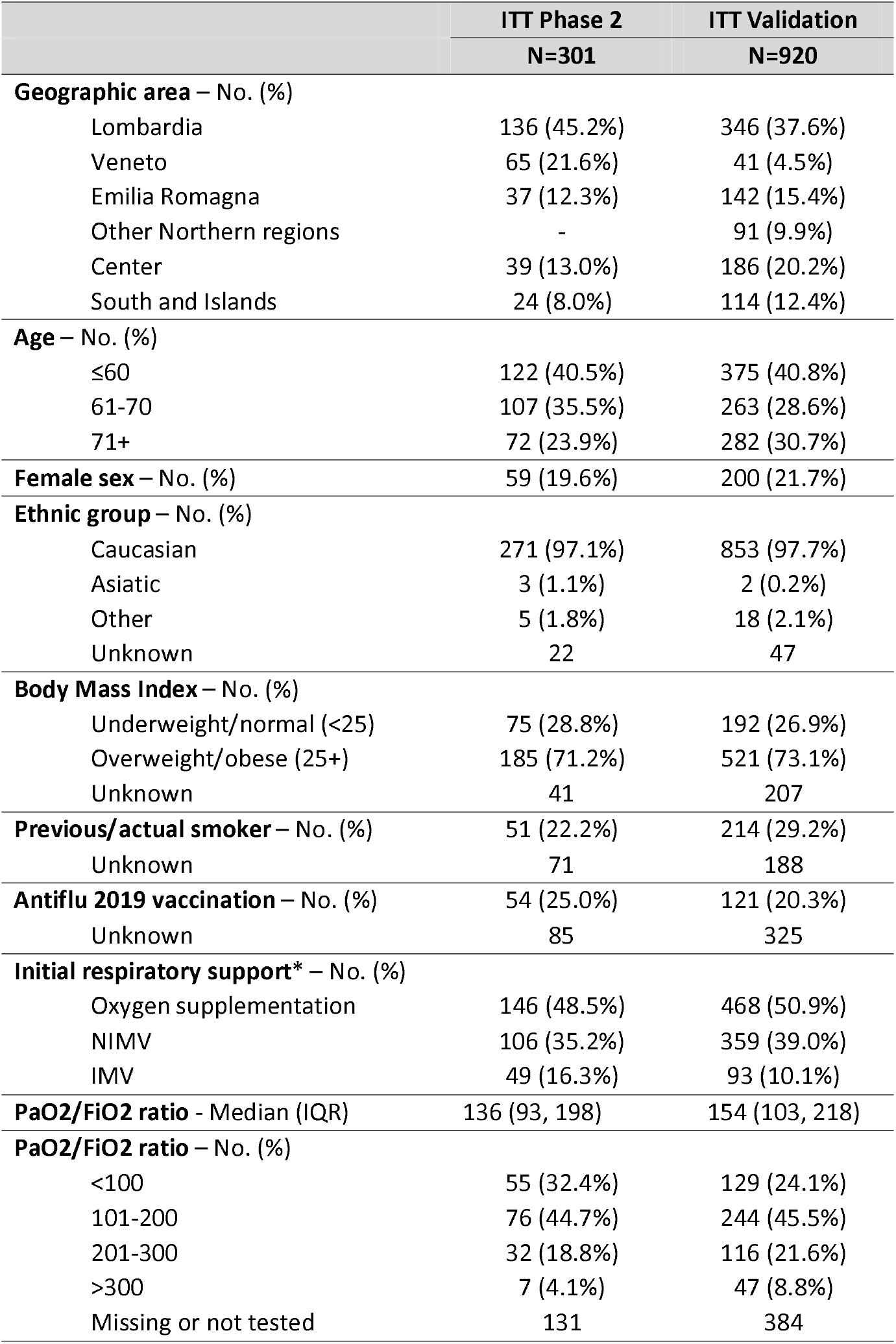
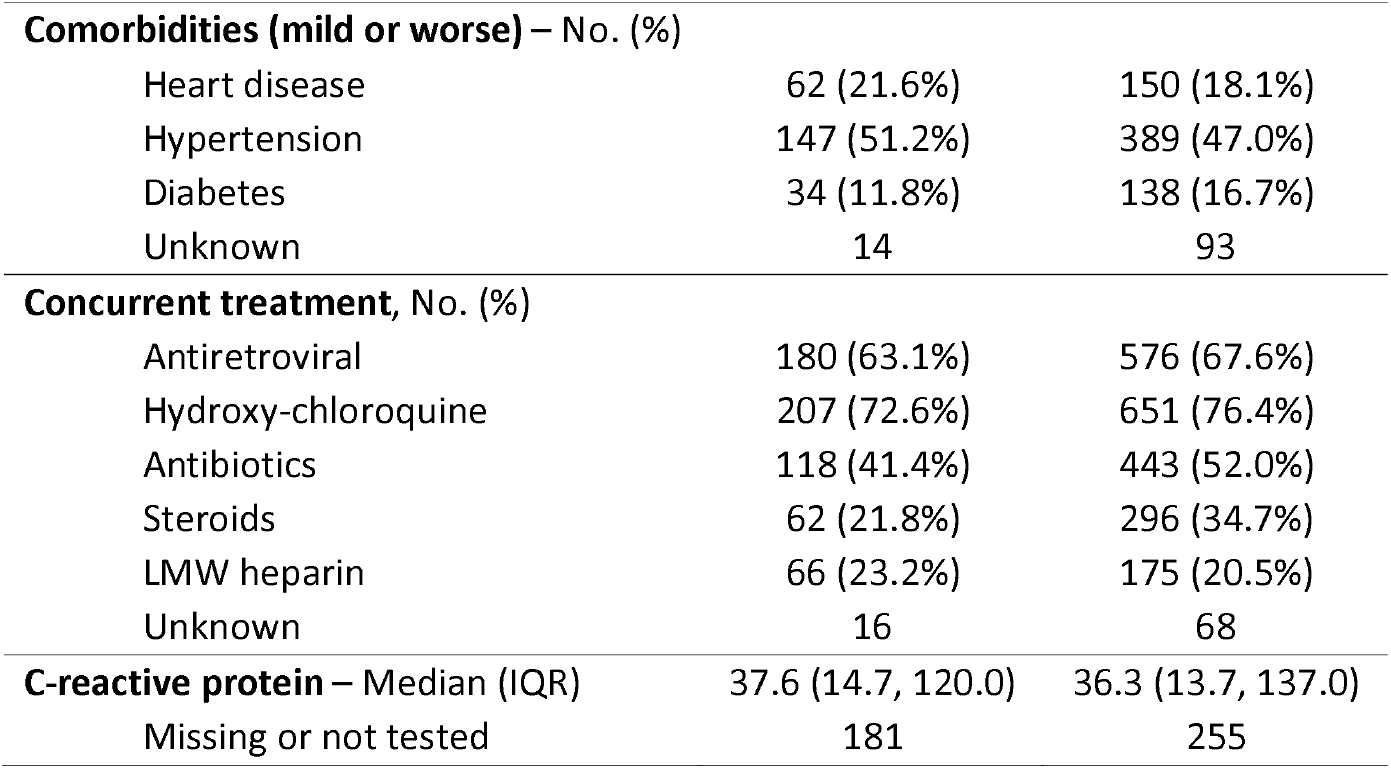
Baseline characteristics of patients in the ITT phase 2 and validation cohorts.

**Figure 1.**
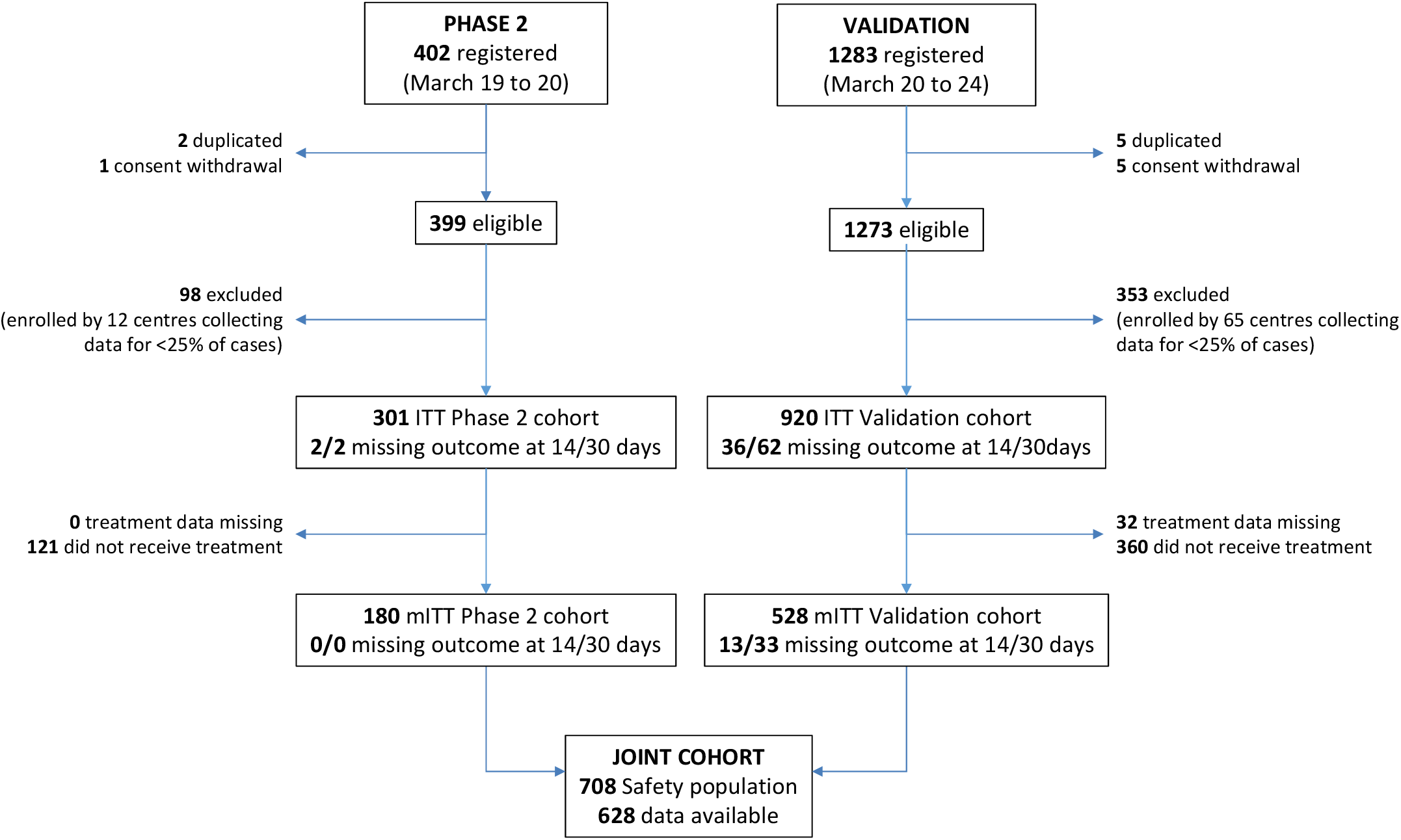
Study flow

Due to lagged drug availability, treatment was given to 59.8% of patients. Median time from registration to treatment administration was 2 days; 23.3% of treated patients received tocilizumab four or more days after registration. The most frequent reason for not giving the drug (once available) was clinical improvement (**eTable 4**, left side). Patients who were younger, and those with worse respiratory function were preferentially treated; also, the geographic location of the centre played a role (**Table 2**, left side).

**Table 2.**
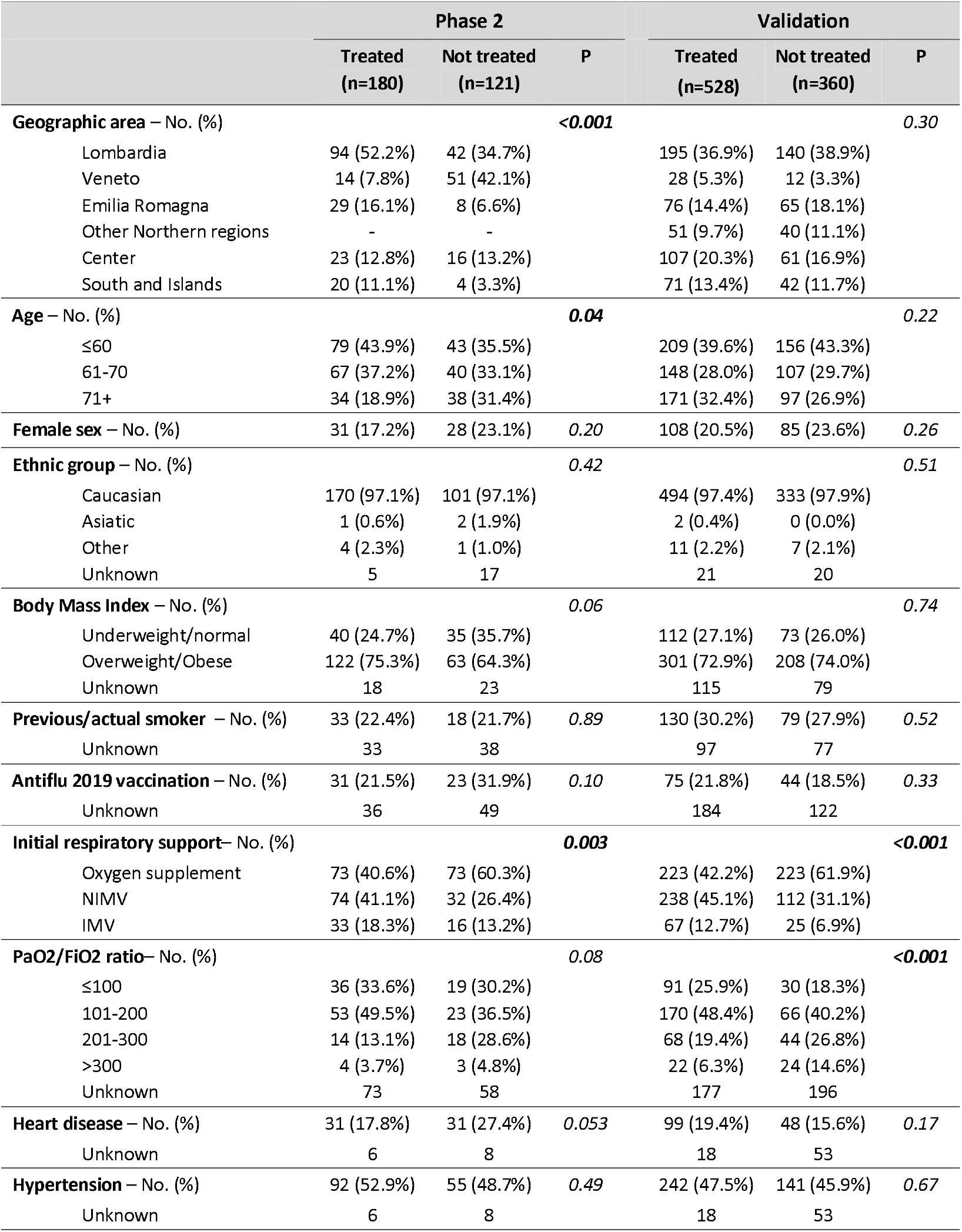
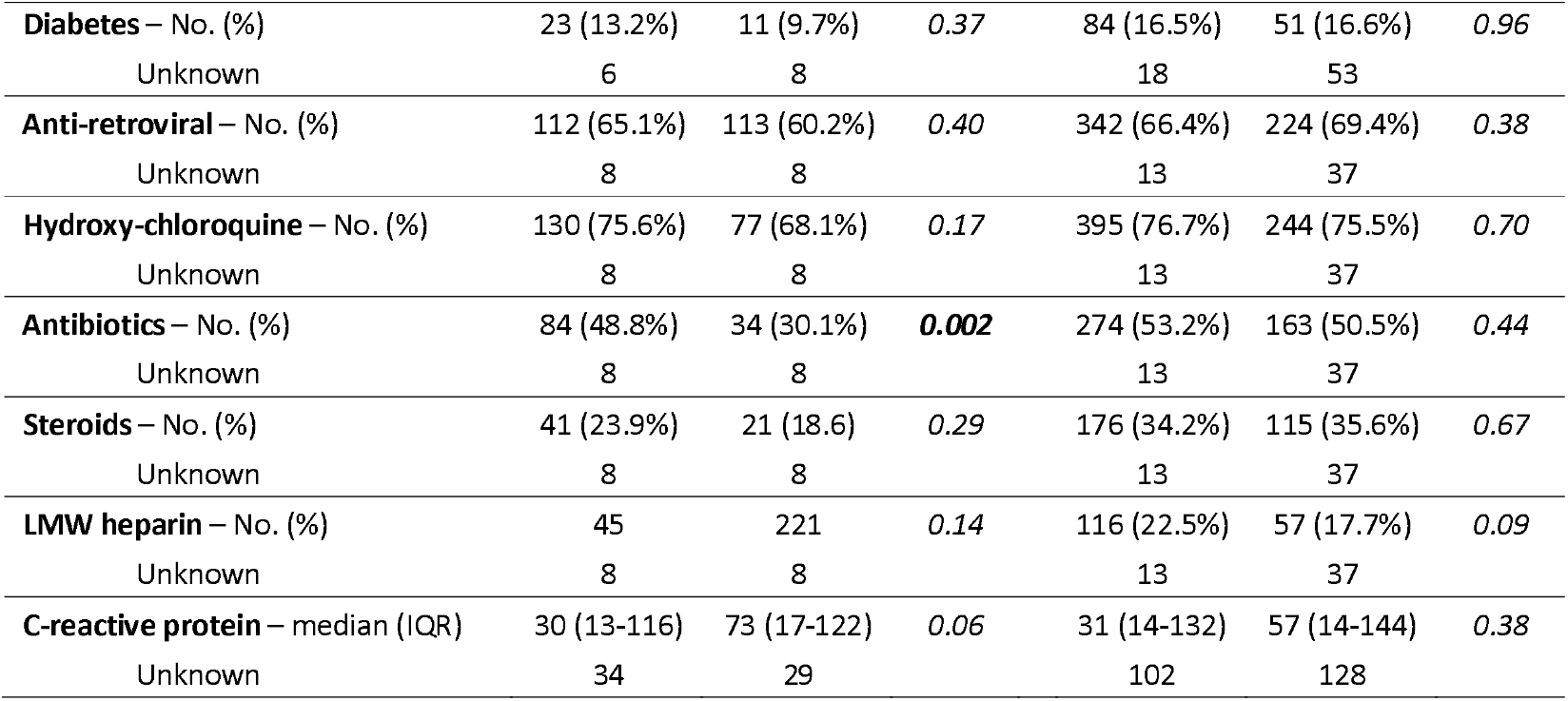
Distribution of baseline characteristics of patients collected at registration by treatment administration.

Overall, 67 (22.3%) deaths were reported in the ITT phase 2 cohort. Lethality rate was 18.4% (97.5% CI: 13.6-24.0) at 14 days and 22.4% (97.5% CI: 17.2-28.3) at 30 days. The null hypothesis was rejected at 30 days but not at 14 days (P<0.001and P=0.52, respectively). At both time points, lethality rates were lower in the mITT population (15.6% and 20.0% -**Table 3**, left side). Due to typical immortal time bias, lethality rates at 14 days were lower for patients receiving treatment four or more days after registration. Risk of death was significantly higher in patients older and with worse PaO2/FiO2 ratio; in addition, lethality rates were lower for patients receiving concurrent corticosteroids, particularly at 14 days where the difference was statistically significant (**Figure 2 and eTable 5**, left side).

**Table 3.**
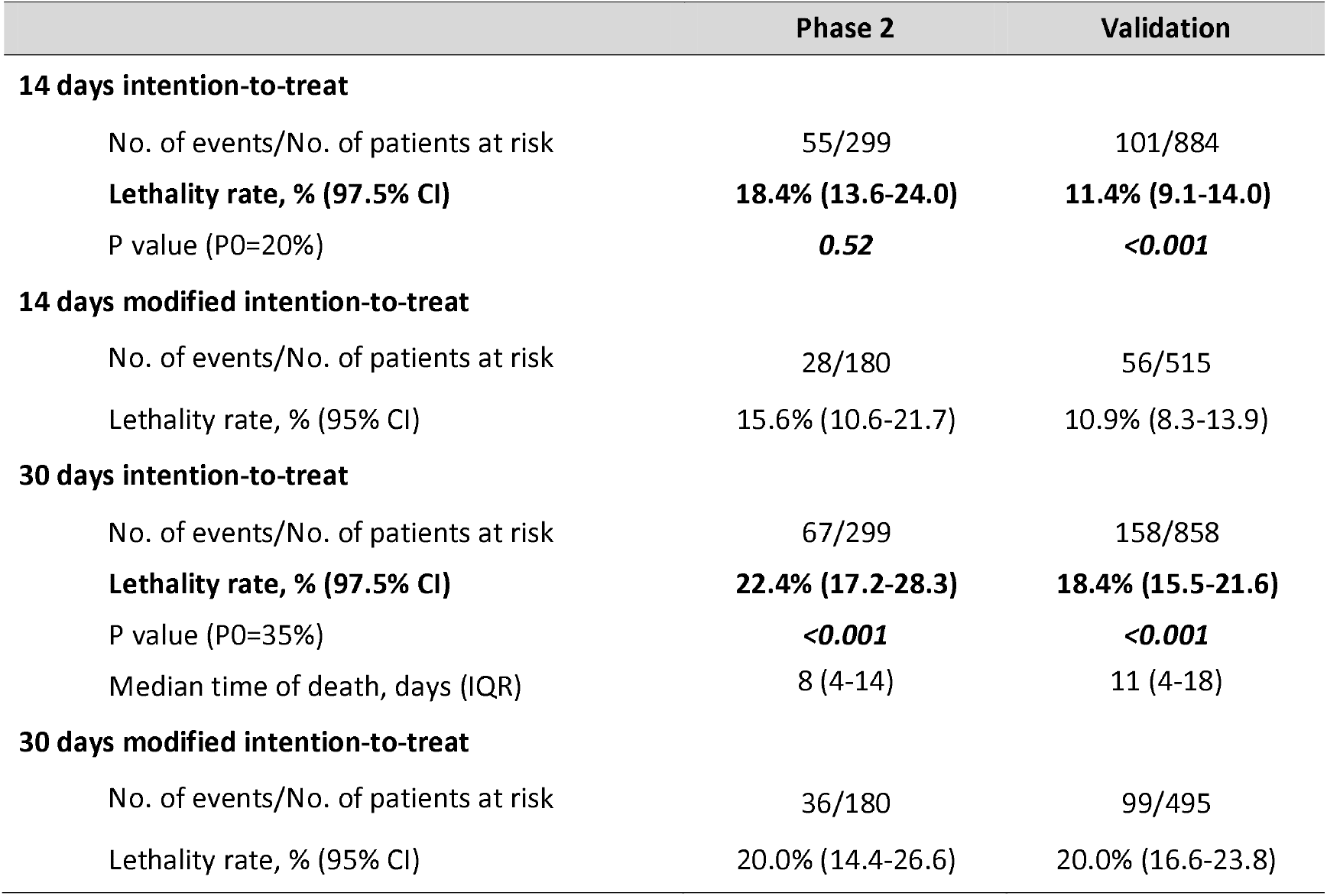
Efficacy analysis.

|  | Phase 2 | Validation |
| --- | --- | --- |
| **14 days intention-to-treat** |  |  |
| No. of events/No. of patients at risk | 55/299 | 101/884 |
| **Lethality rate, % (97.5% CI)** | **18.4% (13.6-24.0)** | **11.4% (9.1-14.0)** |
| P value (P0=20%) | **0.52** | **<0.001** |
| **14 days modified intention-to-treat** |  |  |
| No. of events/No. of patients at risk | 28/180 | 56/515 |
| Lethality rate, % (95% CI) | 15.6% (10.6-21.7) | 10.9% (8.3-13.9) |
| **30 days intention-to-treat** |  |  |
| No. of events/No. of patients at risk | 67/299 | 158/858 |
| **Lethality rate, % (97.5% CI)** | **22.4% (17.2-28.3)** | **18.4% (15.5-21.6)** |
| P value (P0=35%) | **<0.001** | **<0.001** |
| Median time of death, days (IQR) | 8 (4-14) | 11 (4-18) |
| **30 days modified intention-to-treat** |  |  |
| No. of events/No. of patients at risk | 36/180 | 99/495 |
| Lethality rate, % (95% CI) | 20.0% (14.4-26.6) | 20.0% (16.6-23.8) |

**Figure 2.**
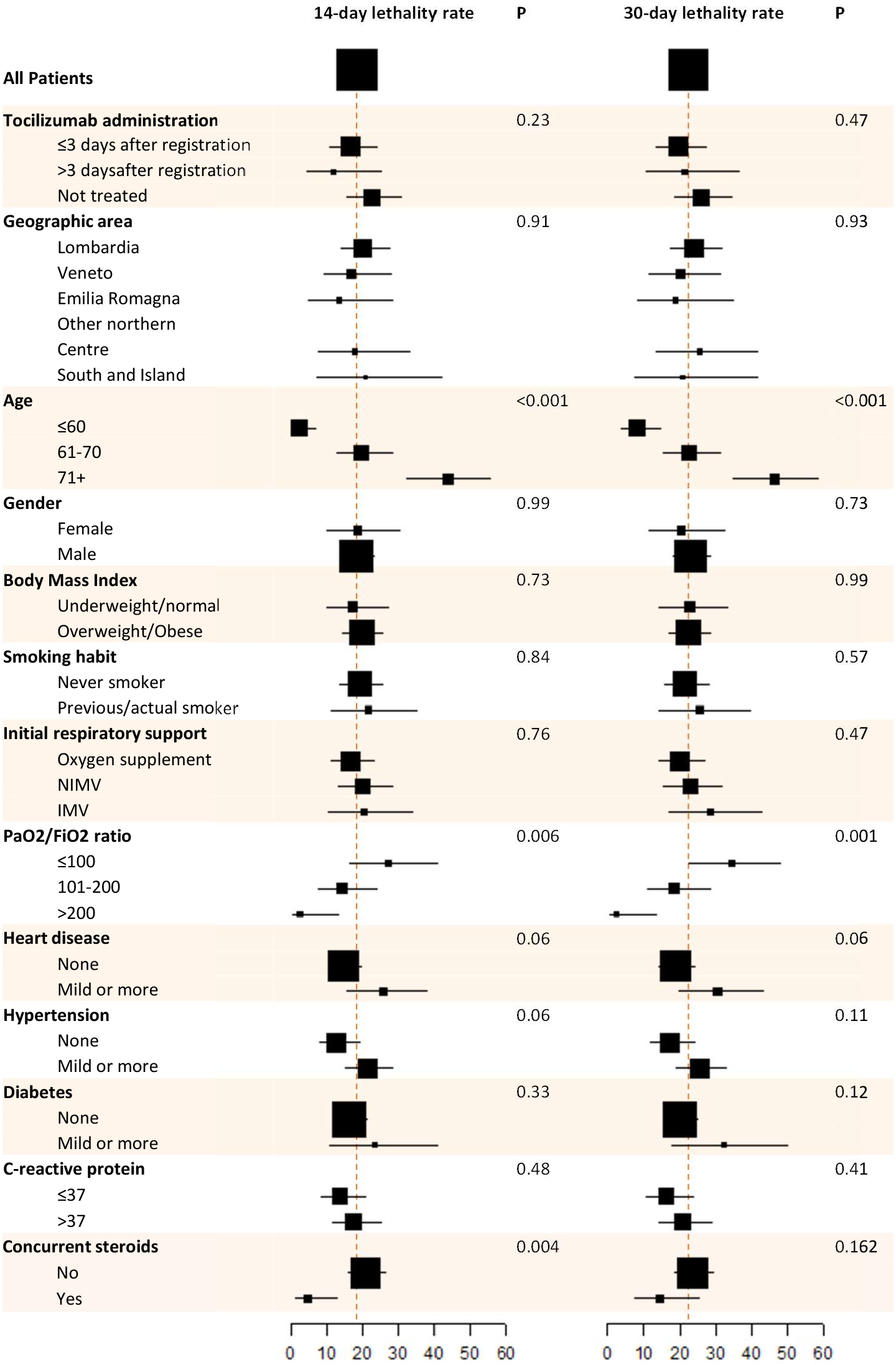
Estimated lethality rates at 14 and 30 days by baseline characteristics of patients in the phase 2 ITT population. Red dash lines represent lethality rates under null hypotheses.

### Validation cohort

The validation cohort included 1273 patients enrolled by 211 centres from March 20^th^ to March 24^th^, 2020 (**Figure 1**, right side). With the same rule applied for phase 2, 65 centres were removed because of missing data, and 920 patients represented the ITT population. Baseline characteristics, shown in tables and figures side by side those of phase 2 patients, were more favorable in the validation than in the phase 2 cohort. Treatment compliance was similar (**eTable 4**, right side). Also in the validation cohort, available treatment was preferentially given to patients with worse respiratory function (**Table 2**, right side). Overall, 158 (17.2%) deaths were reported in the ITT validation cohort. Probability of death was lower in the validation than in the phase 2 cohort, particularly among untreated patients (**eFigure 2**). In the validation cohort, lethality rates were consistently lower than the predefined null hypothesis both at 14 and 30 days in the ITT (11.4% and 18.4%) and mITT (10.9% and 20.0%) populations (**Table 3**, right side). Subgroup analysis of lethality rates produced results similar to those seen in phase 2 (**eFigure 3 and eTable 5**, right side).

### Safety analysis

Safety analysis was done in the joint cohort in 628/708 patients who had received at least one dose of tocilizumab (**eTable 6**). At least one adverse event was reported in 40.8% of patients. Of note, 68 deaths (10.8%) were categorized within adverse events scale.

Causality between such deaths and treatment was described as possible only in one of the 35 cases of respiratory failure. All the other fatal adverse events were reported as unlikely or not related to treatment administration. Seven out of 8 fatal infections were specified as COVID pneumonia. Adverse events that may represent specific side effects of tocilizumab are allergic reactions (3 cases) and ALT or AST increase (reported in 10.5% and 9.1%, respectively) that was severe (grade 3 or 4) in around 3% of cases.

### Hypothesis-generating multivariable analysis

Multivariable analysis was conducted in the joint cohort, after excluding patients treated 4 or more days after registration; 1002 and 980 patients were available for the 14 and 30 days lethality model, respectively (**eTable 7**). Age and respiratory function measured by PaO2/FiO2 ratio were independently significant prognostic factors; the use of corticosteroids was associated with a lower OR of death both at 14 (OR 0.36, 95% CI: 0.21-0.62) and at 30 days (OR 0.62, 95% CI: 0.40-0.95). No significant interaction was found between the effect of tocilizumab and age, gender, PaO2/FiO2 ratio, geographic location and phase 2 vs validation cohorts; also, no interaction was found between the effect of tocilizumab and the use of cortiocosteroids. An interaction was found between treatment and required respiratory support, interaction test p-values being equal to 0.03 and 0.08 at 14 and 30 days, respectively. Specifically, treatment effect on lethality rates was larger among patients not requiring mechanical respiratory support within 24 hours from registration with a OR equal to 0.37 (95% CI: 0.18-0.74) and 0.50 (95% CI: 0.27-0.92) and absolute reductions equal to 7.7% and 6.2%, at 14 and 30 days, respectively (**eFigure 4**).

## Discussion

The primary analysis of the phase 2 TOCIVID-19 study suggests that tocilizumab may reduce lethality at 30 days, although its impact at 14 days seems less relevant. The adverse event profile is consistent with other reports and did not generate clinically relevant warnings, possibly because of the severity of clinical symptoms related to the underlying pathologic condition.[13, 14] Interestingly, the hypothesis-generating multivariable analysis performed to minimize indication and immortal-time biases, showed that the possible effect of tocilizumab might be greater among patients not requiring mechanical ventilation and independent of the effect of corticosteroids; the latter being associated with lower lethality rates, consistently with findings of the Recovery trial. [11] These results support using tocilizumab while waiting for the results of ongoing phase 3 clinical trials. To our knowledge, five ongoing randomised trials are comparing tocilizumab vs placebo (ChiCTR2000029765, NCT04320615, NCT04381936, EudraCT 2020-001408-41, NCT04330638) and another one is comparing immediate vs delayed tocilizumab (NCT04346355). However, some trials have problems in reaching the planned sample size, and most of the trials on medical treatment of COVID-19 are using non validated surrogate outcomes rather than mortality as primary end-point. [15]

TOCIVID-19 is the largest completed prospective study on the effect of tocilizumab using mortality as primary end-point, among published or pre-published reports. Mostly, retrospective or observational data have been reported so far, not based on prospective hypothesis testing, with prevalently positive results.[8, 16-25] However, our study has several limitations that deserve discussion for a better interpretation of findings. The first limitation is the single-arm study design, which prevents definitive conclusions.[26] However, we think that a randomised controlled trial was unfeasible for many reasons. There was a tremendous pressure to have the drug available, due to a widespread media diffusion of positive expectations and the increasing number of patients hospitalized for the disease, as confirmed by the massive registration of centres when the study began. Thus, obtaining a proper informed consent to randomization would have been extremely difficult also due to patients’ condition and clinical burden. Finally, developing a placebo was impossible, and, within a non-blind study, the risk of cross-over from the control to the experimental arm would have been high, reducing the validity of the randomised trial. Within this context, the problem of “learning while doing” was increased.[27] In our opinion, when the TOCIVID-19 trial started this protocol was the best trade-off between do-something and learn-something.

A critical issue of the single-arm design was the definition of the null hypotheses to be tested, already acknowledged in the initial protocol where future modifications of study design were explicitly planned as an option, due to lack of suitable information at the start of the study. Modifying primary end-points while a trial is ongoing is, of course, a risky action. However, we redefined expected benchmarks blind to number and timing of deaths occurring in the study, using data received by an independent research Institution, and looking for a larger absolute benefit as compared to our initial hypothesis.[10] Yet, we cannot be sure that our assumptions are unbiased. A study with data on near 43.000 patients coming from three Italian regions, reports higher lethality at 14 days (22.0%) and lower at 30 days (27.6%) compared to TOCIVID-19 null hypotheses; assuming these estimates as a benchmark, our results would be still clinically significant at both 14 and 30 days.[28]

We tried to take advantage from the availability of a large number of patients registered in the few days immediately following the end of the phase 2 study, identifying a validation cohort. However, even within a very short time window, there was an evident prognostic difference between phase 2 and validation cohorts, the latter having better prognosis than the former.

An operational problem of our study was the discrepancy between timing of drug availability (notwithstanding the commitment of the pharma company) and the extremely high request due to the rapid recruitment rate. As a consequence, two biases arose: the indication bias, occurring when physicians choose to preferentially treat patients with worse prognosis, and the immortal time bias, occurring when treatment administration was delayed as compared to date of registration, since only subjects surviving longer could receive the drug. Actually, the latter bias was particularly evident at 14-day analysis. To minimize both biases, we applied a multivariable logistic regression model, excluding patients receiving the drug later than three days from registration and adjusting for factors affecting the indication of treatment. In addition, we also added concurrent corticosteroid treatment as a confounding variable, following the report of the Recovery trial.[11] Our findings suggest no interaction might exist between the effect of tocilizumab and the concurrent administration of corticosteroids, consistent with another recent report. [29] However, we acknowledge that the comparison between tocilizumab treated and untreated patients, inevitably introduced in the multivariable models, has to be considered explorative and hypothesis-generating because of intrinsic limitations of non-randomised comparison.

Last, we had many missing data, for several reasons: massive involvement and stress of physicians in emergency care; paucity or absence of data-managers; quarantine of paper charts; impracticality of peripheral monitoring; lack of training to the web platform; slow web connections for the study platform due to huge information loading volume. In agreement with IDMC, we tackled the problem by removing un-cooperative centres that provided baseline information for less than 25% of patients; however, we cannot be confident that the remaining missing data are at random.

TOCIVID-19 also has some strengths. As mentioned above, it is the first academic trial promoted in Italy, the largest in terms of centres and patients (being available for the whole Italian territory), assessing a hard endpoint like mortality in a hypothesis-driven design, while off label use of the drug was increasing. [30] In addition, the internal validation, allowed by a companion prospective cohort, contributed to critical interpretation of the results. Further analyses will focus on secondary outcomes (e.g. respiratory outcomes, predictive and prognostic factors, epidemiology insights) and on a larger number of patients.

In conclusion, although with limitations of a phase 2 single arm study, performed in an extremely challenging time and environment, the present study supports the use of tocilizumab, even when corticosteroids are being used, while waiting for results of ongoing phase 3 trials.

## Data Availability

Data will be made available in the near future.

## Authors’ contribution

FP, MCP, PAA, CS, PC, CG designed the study. FP, MCP, GB, CCar, PG, AG, CS managed study conduction. CS, RP, AMM, PP, LF, MMMT, DR, FB, PB, NS, FC, MLM, ML, CCal, NDS, LA, MCa, MCo, GD, NF, FF, MM, VM, CM, EAN enrolled patients and collected study data. LA, PC, CG performed statistical analysis. FP, MCP, PC, CG wrote manuscript draft. All authors have contributed to, seen, and approved the final, submitted version of the manuscript.

## Source of funding and support

No specific funding was available for this study. Tocilizumab was provided by the pharmaceutical company (Roche) free of charge. The National Cancer Institute of Naples was the sponsor of the study and had full property of data and final responsibility for the decision to submit for publication.

## Independent Data Monitoring Committee

An IDMC was appointed on April 9^th^, 2020, including Aldo Maggioni (chief), Paolo Bruzzi, Antonio Pesenti, Valter Torri, and Giuseppe Traversa. Five IDMC meetings were arranged between April 10^th^, 2020 and May 12^th^, 2020. Database was frozen on May 4^th^, 2020.

## Notes

### Competing Interest Statement

Dr. Perrone reports grants, personal fees and non-financial support from Bayer; grants and personal fees from Incyte, Astra Zeneca, Pierre Fabre; personal fees from Celgene, Janssen Cilag, Roche, Pfizer, Sandoz, outside the submitted work.
Dr. Piccirillo reports grants and personal fees from AstraZeneca; grants from Roche; personal fees from Daichii Sankyo, GSK, MSD; non-financial support from Bayer, outside the submitted work.
Dr. Ascierto reports grants and personal fees from BMS, Roche-Genentech, Array; personal fees and other from MSD; personal fees from Novartis, Merck Serono, Pierre Fabre, Incyte, Genmab, NewLink Genetics, Medimmune, AstraZeneca, Syndax, Sun Pharma, Sanofi, Idera, Ultimovacs, Sandoz, Immunocore, 4SC, Alkermes, Italfarmaco, Nektar, Boehringer-Ingelheim, outside the submitted work.
Dr. Salvarani reports grants and personal fees from Roche; personal fees from Sanofi-Genzyme, Abbvie, Pfizer, Eli-Lilly, Novartis, outside the submitted work.
Dr. Castelli reports grants from Gilead, ViiV, GSK, Janssen, Eiger, Roche, Gilead, outside the submitted work.
Dr. Gravina reports non-financial support from Pfizer, outside the submitted work.
Dr. Lichtner reports grants from Gilead; personal fees from Abbvie, Merck, Janseen, Angelini, outside the submitted work.

### Clinical Trial

NCT04317092

### Clinical Protocols

http://www.repo.epiprev.it/1604

http://www.repo.epiprev.it/1610

### Author Declarations

According to a specific Italian law, TOCIVID-19 was approved for all Italian centers by the National Ethical Committee at the Lazzaro Spallanzani Institute on March 18th, 2020; two amendments followed on March 24th, 2020 and April 28th, 2020.

## References

1. Berlin DA, Gulick RM, Martinez FJ. Severe Covid-19. N Engl J Med 2020.

2. Grasselli G, Zangrillo A, Zanella A, et al. Baseline Characteristics and Outcomes of 1591 Patients Infected With SARS-CoV-2 Admitted to ICUs of the Lombardy Region, Italy. JAMA 2020.

3. Guan WJ, Ni ZY, Hu Y, et al. Clinical Characteristics of Coronavirus Disease 2019 in China. N Engl J Med 2020; 382:1708–20.

4. Huang C, Wang Y, Li X, et al. Clinical features of patients infected with 2019 novel coronavirus in Wuhan, China. Lancet 2020; 395:497–506.

5. Zhu N, Zhang D, Wang W, et al. A Novel Coronavirus from Patients with Pneumonia in China, 2019. N Engl J Med 2020; 382:727–33.

6. Le RQ, Li L, Yuan W, et al. FDA Approval Summary: Tocilizumab for Treatment of Chimeric Antigen Receptor T Cell-Induced Severe or Life-Threatening Cytokine Release Syndrome. Oncologist 2018; 23:943–7.

7. Scott LJ. Tocilizumab: A Review in Rheumatoid Arthritis. Drugs 2017; 77:1865–79.

8. Xu X, Han M, Li T, et al. Effective treatment of severe COVID-19 patients with tocilizumab. Proc Natl Acad Sci U S A 2020.

9. Piccirillo M, Ascierto P, Atripaldi L, et al. Multicenter study on the efficacy and tolerability of tocilizumab in the treatment of patients with COVID-19 pneumonia. Epidemiologia & Prevenzione 2020:www.repo.epiprev.it/1604

10. Chiodini P, Arenare L, Piccirillo M, Perrone F, Gallo C. A phase 2, open label, multicenter, single arm study of tocilizumabon the efficacy and tolerability of tocilizumab in the treatment of patients with COVID-19 pneumonia (TOCIVID-19 trial): Statistical Analysis Plan. Epidemiologia & Prevenzione 2020:www.repo.epiprev.it/1610.

11. Horby P, Lim WS, Emberson J, et al. Effect of Dexamethasone in Hospitalized Patients with COVID-19: Preliminary Report. 2020:2020.06.22.20137273.

12. Agresti A, Caffo B. Simple and Effective Confidence Intervals for Proportions and Differences of Proportions Result from Adding Two Successes and Two Failures. The American Statistician 2000; 54:280-8.

13. Giacobbe DR, Battaglini D, Ball L, et al. Bloodstream infections in critically ill patients with COVID-19. European journal of clinical investigation 2020:e13319.

14. Piano S, Dalbeni A, Vettore E, et al. Abnormal liver function tests predict transfer to intensive care unit and death in COVID-19. Liver international : official journal of the International Association for the Study of the Liver 2020.

15. Desai A, Gyawali B. Endpoints used in phase III randomized controlled trials of treatment options for COVID-19. EClinicalMedicine 2020; 23:100403.

16. Alattar R, Ibrahim TBH, Shaar SH, et al. Tocilizumab for the treatment of severe coronavirus disease 2019. J Med Virol 2020.

17. Antwi-Amoabeng D, Kanji Z, Ford B, Beutler BD, Riddle MS, Siddiqui F. Clinical outcomes in COVID-19 patients treated with tocilizumab: An individual patient data systematic review. J Med Virol 2020.

18. Capra R, De Rossi N, Mattioli F, et al. Impact of low dose tocilizumab on mortality rate in patients with COVID-19 related pneumonia. Eur J Intern Med 2020.

19. Colaneri M, Bogliolo L, Valsecchi P, et al. Tocilizumab for Treatment of Severe COVID-19 Patients: Preliminary Results from SMAtteo COvid19 REgistry (SMACORE). Microorganisms 2020; 8.

20. Guaraldi G, Meschiari M, Cozzi-Lepri A, et al. Tocilizumab in patients with severe COVID-19: a retrospective cohort study. The Lancet Rheumatology 2020.

21. Jordan SC, Zakowski P, Tran HP, et al. Compassionate Use of Tocilizumab for Treatment of SARS-CoV-2 Pneumonia. Clinical infectious diseases : an official publication of the Infectious Diseases Society of America 2020.

22. Klopfenstein T, Zayet S, Lohse A, et al. Tocilizumab therapy reduced intensive care unit admissions and/or mortality in COVID-19 patients. Med Mal Infect 2020.

23. Rojas-Marte GR, Khalid M, Mukhtar O, et al. Outcomes in Patients with Severe COVID-19 Disease Treated with Tocilizumab - A Case-Controlled Study. QJM : monthly journal of the Association of Physicians 2020.

24. Sciascia S, Aprà F, Baffa A, et al. Pilot prospective open, single-arm multicentre study on off-label use of tocilizumab in patients with severe COVID-19. Clinical and experimental rheumatology 2020; 38:529–32.

25. Toniati P, Piva S, Cattalini M, et al. Tocilizumab for the treatment of severe COVID-19 pneumonia with hyperinflammatory syndrome and acute respiratory failure: A single center study of 100 patients in Brescia, Italy. Autoimmun Rev 2020:102568.

26. Goodman JL, Borio L. Finding Effective Treatments for COVID-19: Scientific Integrity and Public Confidence in a Time of Crisis. JAMA 2020.

27. Angus DC. Optimizing the Trade-off Between Learning and Doing in a Pandemic. JAMA 2020.

28. Giorgi Rossi P, Ferroni E, Spila Alegiani S, et al. Survival of hospitalized COVID-19 patients in Northern Italy: a population-based cohort study by the ITA-COVID19 Network. MedrXiv 2020:2020.05.15.20103119.

29. Sanz Herrero F, Puchades Gimeno F, Ortega García P, Ferrer Gómez C, Ocete Mochón MD, García Deltoro M. Methylprednisolone added to tocilizumab reduces mortality in SARS-CoV-2 pneumonia: An observational study. n/a.

30. Addis A, Genazzani A, Trotta MP, Magrini N. Promoting Better Clinical Trials and Drug Information as Public Health Interventions for the COVID-19 Emergency in Italy. Ann Intern Med 2020.

